# Chronic prescription opioid use in MS: Associations with adverse outcomes and acute medical service utilization

**DOI:** 10.1101/2025.09.09.25335375

**Authors:** Aaron P. Turner, Carol Malte, Anne Arewasikporn, Eric J. Hawkins, Pradeep Suri, Stephen P. Burns, Steven L. Leipertz, Jodie K. Haselkorn

## Abstract

**Background:** The purpose of this study is to examine whether chronic prescription opioid use in individuals with MS is associated with adverse medical outcomes and acute medical service utilization.

**Methods:** This propensity score-matched, retrospective, cohort study included data from 15,377 Veterans with MS obtained from national VA administrative data. Veterans with filled prescriptions for chronic opioid therapy (*n*=1,908) in 2017 were matched 1:1 and compared to those not prescribed chronic opioid therapy using logistic regression models adjusted for propensity score, age, sex, race, and ethnicity. Outcomes included falls, fractures, wounds, and all-cause mortality, as well as acute inpatient and emergency department utilization, and intensive care unit admission in 2018.

**Results:** The chronic opioid group had a higher likelihood of a fall (OR_adj_ =1.77, 95% CI, 1.03– 3.02, *p*=0.038), inpatient admission (OR_adj_=1.21, 95% CI, 1.02–1.43, *p*=0.028), and emergency department visit (OR_adj_=1.25, 95% CI, 1.09–1.43, *p*=0.001). Significant differences were not detected between those prescribed and not prescribed chronic opioid therapy on fractures, wounds, all-cause mortality, and intensive care unit admission, although in all cases the occurrence of these outcomes was higher in the opioid group.

**Conclusion:** Chronic prescription opioid use among Veterans with MS was associated with a higher likelihood of adverse outcome (falls) and higher intensity health care utilization (inpatient admissions and emergency department visits) in the following year. Education regarding MS-specific risks of opioid use should be provided at initial prescription and re-evaluation of risks versus benefits should be conducted regularly.

## 1.0 Introduction

Pain is a significant problem for people with MS. At any given time, it is estimated that roughly half will report pain that has a noticeable impact on their lives and as many as one third will report pain that is chronic (lasting 3 months or greater) and severe.^1-3^ MS-related pain can have varying presentations (e.g., neuropathic, musculoskeletal, headache)^4,5^ and locations (e.g., extremities, back, jaw),^6^ and can negatively impact functioning and quality of life.^7,8^

Given the frequent occurrence of pain in MS it is unsurprising that both acute and chronic prescription opioid use (defined as <90 days and ≥90 days respectively), remain common.^9^ Although current evidence suggests that the long-term efficacy of opioids for chronic pain is limited and side effects are frequent,^10^ many individuals with MS continue to receive prescribed opioids for years. For example, in one recent examination of chronic prescription opioid use among individuals with MS, over a three-year period approximately 97% of those with chronic prescription use had chronic use in the prior year (less than 3% was new chronic prescription opioid use in any given year).^9^

In general population samples, chronic prescription opioid use is associated with several adverse outcomes including increased risk of falls, fractures, and mortality.^11-16^ It is also associated with greater use of acute medical services including emergency department (ED) visits and hospital admissions among individuals with common chronic pain complaints such as low back pain.^17^ There is substantial need to understand the risks of adverse outcomes of chronic prescription opioid use in people with MS.

The primary aim of this study was to examine whether chronic prescription opioid use (versus no chronic use) increases the likelihood of adverse outcomes and acute medical service utilization in Veterans with MS.

## 2.0 Method

### 2.1 Participants

U.S. Veterans who had received services in the Veterans Health Administration between 2016 and 2018 and had a confirmed MS diagnosis prior to 2017 (*N* = 15,377) were identified from the VA Corporate Data Warehouse (CDW), a repository of information derived from the VA’s national electronic medical record. Inclusion and exclusion criteria for a MS diagnosis were defined as any combination of ≥ 3 MS-related inpatient or outpatient encounters or MS disease modifying therapy prescriptions in a 1-year period, which have been shown to reduce case-finding bias.^18^ Similar MS case finding methodology has been used in prior studies.^7,9,19-26^

Veterans who were deceased prior to 2018, received methadone or buprenorphine for opioid use disorder treatment in 2016-2017, or were actively enrolled in cancer or hospice care in 2016-2017 were excluded. This study was approved by the human subjects committee of the VA Puget Sound Health Care System. Written consent was waived for this project.

### 2.2 Measures

#### 2.2.1 Demographics

All demographic data, including age, sex, race, ethnicity, marital status and rural designation, were obtained from the CDW. Rurality was defined using geographic information systems mapping of individual addresses to US Census tracts, which were then coded using

U.S. Department of Agriculture Rural-Urban Commuting Area (RUCA) designations collapsed into categories of urban vs. rural.^27^ Demographic variables are listed in Table 1 based on how they were used for propensity matching (described below).

**Table 1.**
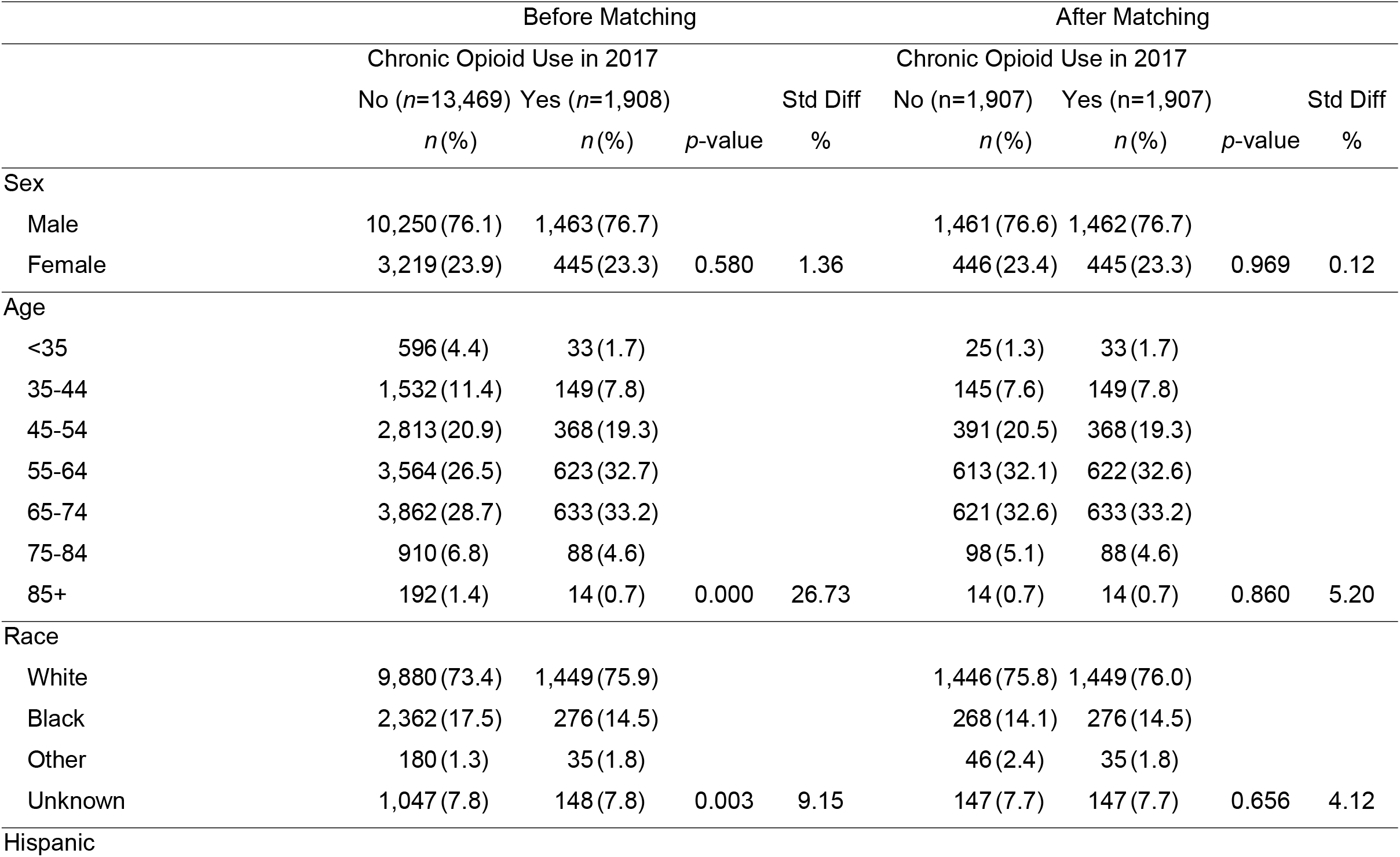
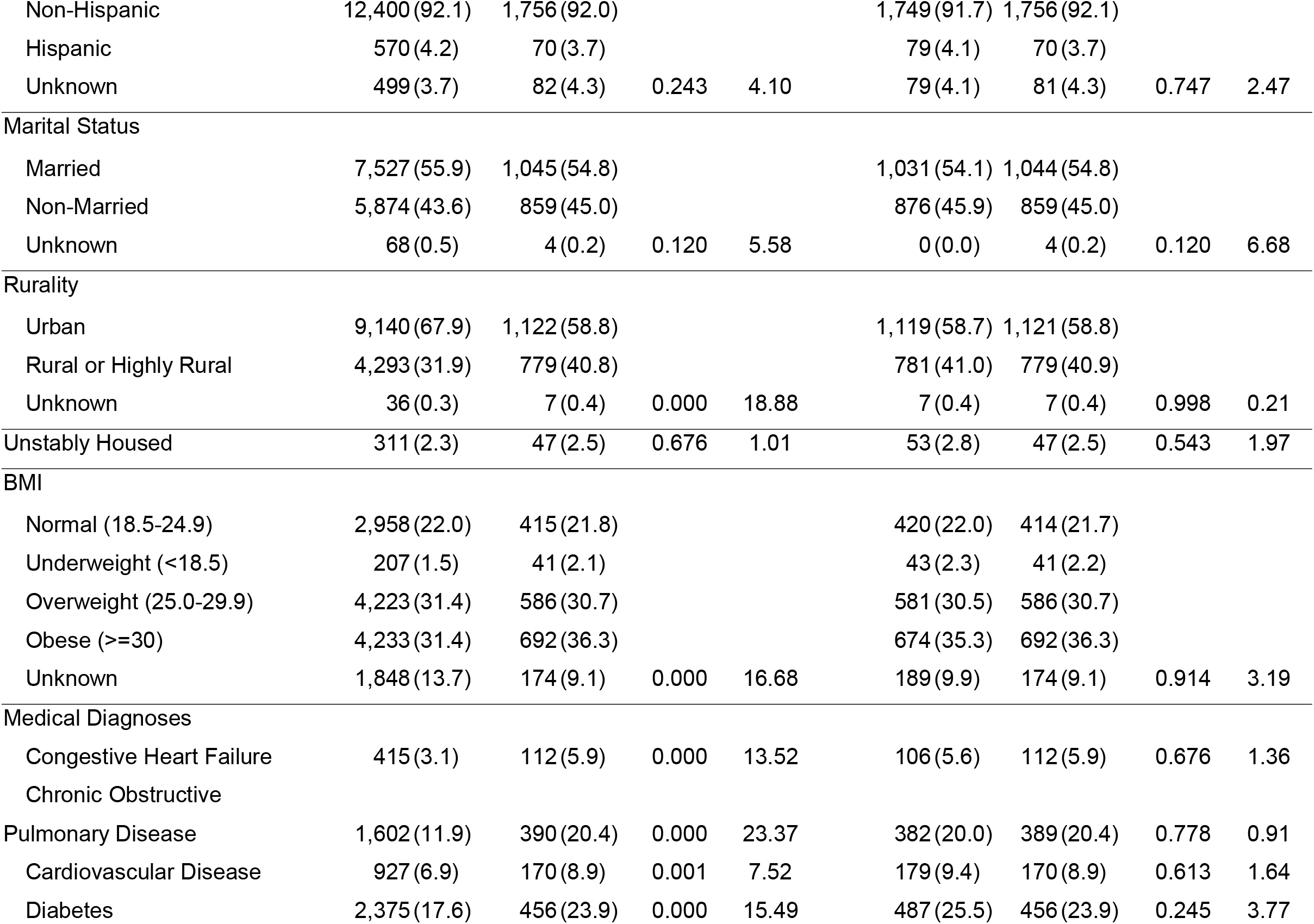
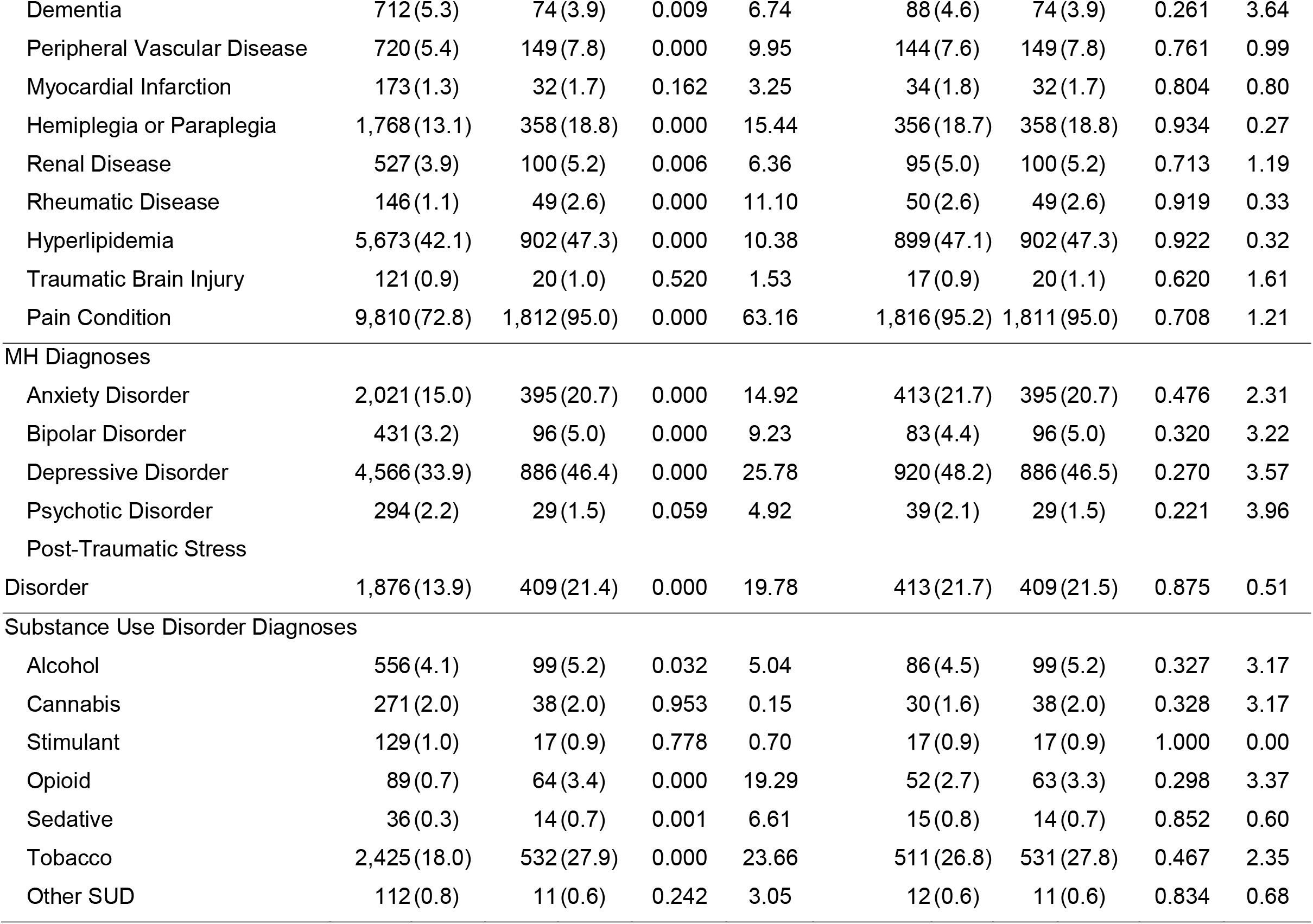
2016-2017 Demographics and Clinical Characteristics Before and After Matching

### 2.2.2 Comorbidities

Medical, psychiatric, and substance use diagnoses included in propensity models are listed in Table 1. Comorbidities were coded based upon any occurrence in 2016-2017 as 1 = present, 0 = absent. The International Classification of Diseases, Tenth Revision Codes (ICD-10) were used to construct variables identifying comorbidities according to a classification system utilized in previous studies to examine opioid use in Veterans.^28^ Pain diagnoses were identified and categorized according to an established pain classification system for identifying 12 common pain conditions using ICD codes.^29^ Pain was similarly coded as 1 = present, 0 = absent for Veterans meeting criteria for any of the 12 conditions in 2016-2017.

#### 2.2.3 Chronic Prescription Opioid Use

Prescription opioid medication data were obtained from outpatient pharmacy files in the CDW that included medication type, dose, days supplied and release date. Opioid medications included codeine, morphine, oxycodone, hydrocodone, oxymorphone, propoxyphene, hydromorphone, levorphanol, meperidine, methadone, tramadol, pentazocine, and fentanyl.

Total days’ supply was obtained by summing the days’ supply from all opioid prescriptions filled during 2017. Measures of continuous supply were calculated as a proxy for opioid use using the ‘as prescribed’ approach, which assumes patients take medications at the prescribed dose and schedule. Chronic opioid therapy was defined as having a filled prescription for ≥ 90 days continuous supply of any previously described opioid medication and no chronic opioid therapy was defined as < 90 days continuous supply (including no opioid use). For simplicity, filled prescriptions for opioids are described as prescribed opioid use or prescribed opioid therapy.

#### 2.2.4 Outcomes

Data about adverse medical outcomes in calendar year 2018 were obtained from the CDW and included information about falls, fractures, wounds and all-cause mortality. ICD-10 codes were used to construct variables for falls (W00-W19), fractures (S02, S12, S22,…S92, M80, M48, M84 [excluding concussion, dental, fatigue, sequelae]), and wounds (L89). Mortality was derived from the VA vital status file, which combines mortality data from multiple VA and non-VA sources.

Acute medical service utilization data were also extracted from the CDW and included new acute inpatient hospitalizations, intensive care unit (ICU) admissions and ER visits. Inpatient hospitalizations were identified by CDW inpatient admission table data. ICU admission and ER visits were identified using algorithms created by the VA Informatics and Computing Infrastructure (VINCI) to identify these services from key words extracted from clinic identification codes (stop codes) and clinic names.

### 2.3 Matching

To reduce potential confounding, Veterans with MS prescribed chronic opioid therapy in 2017 were matched to Veterans not prescribed chronic opioid therapy according to propensity score, representing the predicted probability of a Veteran receiving chronic opioid therapy and derived from a logistic regression model that included 34 covariates. Propensity scores were matched 1:1 using a “greedy” matching algorithm^30^ and a maximum caliper of 0.1. All variables in Table 1 were used to calculate propensity scores.

### 2.4 Statistical Analysis

First, descriptive statistics were calculated to characterize overall sample and verify assumptions necessary for analysis were met.

The effectiveness of propensity score matching was evaluated by comparing Veterans prescribed chronic opioid therapy in 2017 to matched Veterans not prescribed chronic opioid therapy on all baseline clinical characteristics included in the propensity model using chi-square tests and standard mean differences. The match was considered successful if standard mean differences were <10%.

Primary analyses compared the adjusted likelihood of each of the four adverse medical outcomes and three service utilization outcomes specified above during 2018 between patients receiving chronic prescription opioid therapy in 2017 and matched controls not receiving chronic prescription opioid therapy during the same period using logistic regression. Analyses were structured to ensure that the exposure (chronic opioid prescription) preceded outcome to reduce the potential for reciprocal effects (e.g., a fall or fracture resulted in a subsequent new opioid prescription). Given that prior analyses had established that incident chronic opioid use was extremely low in this population (initiation of chronic opioid prescribing occurred in less than 3% of participants per year)^9^ a time-to-event strategy was not adopted. Logistic models included an indicator of chronic opioid use (0/1) and were adjusted for baseline propensity score, continuous age, sex (male/female), race (White, Black, other race, unknown race) and ethnicity (Hispanic, Non-Hispanic, unknown ethnicity).

## 3.0 Results

### 3.1 Sample Characteristics

Prior to matching, participants in the sample (n=15,377) were on average, male (76.2%), middle-aged (*M*=58.6, *SD*=12.7), and of non-Hispanic (92.1) descent. The majority were married (55.7%) and resided in an urban area (66.7%).

### 3.2 Propensity Score Matching

Of the 15,377 Veterans eligible for study inclusion, 1,908 (12.4%) received chronic opioids in 2017. Prior to matching, those prescribed and not prescribed chronic opioid therapy differed on baseline characteristics, with standardized differences exceeding 10% for a number of covariates, with the greatest standardized differences (≥20%) seen in age and pain condition, depressive disorder, tobacco use disorder, chronic obstructive pulmonary disease and post-traumatic stress disorder diagnoses (Table 1). After propensity score matching, 1,907 Veterans receiving chronic opioid therapy were matched to 1,907 who were not. The matched groups were comparable on demographic and clinical variables, with most standard differences less than 2.5% and no differences exceeding 6.7% (Table 1). The matched sample overall was 62.0 years old (*SD* = 10.9) and 23.4% were women (Table 1). Pain disorders (95.1%), hyperlipidemia (47.2%) and diabetes (24.7%) were the most common medical comorbidities, depressive disorders (47.4%) and post-traumatic stress disorder (21.6%) were the most common psychiatric comorbidities and tobacco use disorder was the most common substance use disorder (27.3%).

### 3.3 Adverse Medical Outcomes and Acute Service Utilization

With respect to adverse medical outcomes, in 2018 there was a higher likelihood of falls among those prescribed chronic opioid therapy in 2017 (1.9% vs. 1.2%; OR_adj_=1.77; 95% CI, 1.03, 3.02; *p*=0.038). No statistically significant differences were seen in the likelihood of fractures or wounds between matched cohorts. In 2018, Veterans prescribed chronic opioid therapy in 2017 experienced 61 (3.2%) deaths, relative to 51 (2.7%) among the matched sample not prescribed chronic opioid therapy. However, after adjusting for propensity score, age, sex and race/ethnicity, we did not detect differences in the likelihood of death in 2018. Notably, the frequencies of all adverse medical outcomes were higher in the chronic opioid group, even when the between-group differences were not statistically significant (Table 2).

**Table 2.** 2018 Outcomes by 2017 Chronic Opioid Receipt

|  | Chronic Opioid Use in 2017 |  | OR* | 95% CI | *p*-value |
| --- | --- | --- | --- | --- | --- |
|  | No<br>*n* (%) | Yes<br>*n* (%) |  |  |  |
| Adverse Outcome |  |  |  |  |  |
| Death | 51 (2.7) | 61 (3.2) | 1.24 | (0.84 - 1.81) | 0.275 |
| Wound | 110 (5.8) | 114 (6.0) | 1.05 | (0.80 - 1.38) | 0.736 |
| Fall | 22 (1.2) | 37 (1.9) | 1.77 | (1.03 - 3.02) | 0.038 |
| Fracture | 41 (2.2) | 52 (2.7) | 1.29 | (0.85 - 1.95) | 0.235 |
| Acute Medical Service Utilization |  |  |  |  |  |
| ER Visit | 652 (34.2) | 749 (39.3) | 1.25 | (1.09 - 1.43) | 0.001 |
| Inpatient Admission | 311 (16.3) | 362 (19.0) | 1.21 | (1.02 - 1.43) | 0.028 |
| ICU Admission | 18 (0.9) | 24 (1.3) | 1.34 | (0.72 - 2.48) | 0.352 |

\*Adjusted for propensity score, continuous age, sex, race, and ethnicity.
Total N for each group = 1907.

With respect to acute service utilization outcomes, significant differences were seen in those prescribed and not prescribed chronic opioid therapy with respect to emergency room visits (39.3% vs. 34.2%; OR_adj_=1.25; 95% CI, 1.09, 1.43; *p*=0.001) and inpatient admissions (19.0% vs. 16.3%; OR_adj_=1.21; 95% CI, 1.02, 1.43; *p*=0.028). Rates of ICU admissions were slightly higher among those prescribed chronic opioid therapy relative to those who were not but the difference between rates was not significant. (Table 2).

## 4.0 Discussion

This study examined outcomes associated with chronic prescription opioid use among individuals with MS using administrative data from a large national healthcare system (U.S. Department of Veterans Affairs). Propensity score matching with multivariable adjustment was used to compare individuals with and without a chronic opioid prescription and to identify differences in adverse medical events and acute service utilization in the following year.

With respect to adverse medical outcomes, chronic prescription opioid use was associated with an increased likelihood of falls among individuals with MS. This finding is consistent with several recent review articles examining opioid use among general population samples of older adults.^15,16^ Drowsiness, orthostatic hypotension, and hyponatremia resulting from opioid use have all been proposed as potential causal mechanisms.^15^ Falls are also common for individuals with MS, with some studies reporting annual fall rates of 50% or greater.^31^ Fall risk in MS has been attributed to slowed somatosensory conduction as well as altered central integration.^31^ Thus, it is likely that reduced mobility common in both groups (MS and older adults) is a common risk factor that may be exacerbated by chronic prescription opioid use. Future studies should examine the interaction between mobility, opioid use and falls in individuals with MS. Particular attention should be paid to periods of declining mobility immediately preceding the adoption of higher level ambulatory assistive devices (e.g., the timeframe prior to the adoption of a cane) when the risk of falls is higher.

The absence of significant association between chronic prescription opioid use and fractures in this sample is surprising given the frequency of falls in individuals with MS^31^ as well as the relative increase in those receiving chronic opioid prescription in MS found in this study. It is also inconsistent with several recent meta analyses examining opioid use among older adults more generally.^16^ Similarly, all-cause mortality has been associated with opioid use in other studies,^11-14^ but not in the current investigation. The proposed association between chronic opioid prescription and wounds was also not supported, though this hypothesis was exploratory. Although not statistically significant in every case and therefore not reliable, it is worthy of note that for all outcomes examined, the likelihood of adverse medical outcomes increased with chronic opioid use. Such relationships might be more readily apparent in studies with greater power or longer follow-up assessment.

With respect to acute service utilization, chronic prescription opioid use was associated with an increased likelihood of ER visit or inpatient admission the following year. This finding is consistent with literature in other populations such as low back pain, in which prescription opioid use, particularly but not exclusively chronic use, has been linked to a higher rate of clinic visits, as well as a greater likelihood of an ER visit or hospital admission.^17^

Study findings highlight the importance of considering the consequences of chronic opioid prescription both in terms of adverse outcomes and increased acute service utilization. Shared decision making between patients and providers should consider both benefits and risks, as well as available non-pharmacologic strategies for pain management with demonstrated efficacy in MS (including but not limited to transcutaneous electrical nerve stimulation, Ai Chi exercise, hypnosis)^32,33^ and chronic pain more generally (mindfulness meditation, cognitive behavioral therapy).^34,35^ Ongoing use of opioids should be reviewed at regular intervals.

### Limitations

This study has several strengths worthy of note. It utilizes medical record data from the U.S. Department of Veterans Affairs, reflecting a large, geographically diverse population of individuals with MS from the largest integrated health care service in the United States. The sample contains a relatively large proportion of men, who are frequently under-represented in MS research. At the same time, medical record data brings well known limitations. Opioid use was estimated via proxy (filled prescriptions) and it is possible that some prescriptions, despite being filled, were never actually used, or that the medication was not taken exactly as prescribed. The regulation and monitoring of opioid medications (e.g., controlled substance status, monitoring via the VA Opioid Safety Initiative) mitigates this concern relative to many other medications. Study methodology did not allow for examination of non-prescribed opioid use or opioid prescriptions from outside the VA. Estimates for the rate of falls and fractures rely on coding of such events in the medical record, and likely under-identify these events relative to self-report.^31^ However, those that were coded likely represent events with more serious injuries. Fracture events are likely underestimated in both groups since fracture care may have been received outside of the VA. The VA medical record and VA CDW do not include information required to characterize MS disease process such as MS severity or subtype. As with all observational studies, results could be impacted by unmeasured confounding. Finally, findings may not generalize to Veterans who do not receive their care at VA or non-Veterans receiving care in other medical settings.

## 5.0 Conclusion

Chronic prescription opioid use among Veterans with MS was associated with adverse outcomes (falls) and greater use of acute medical services (inpatient admissions and emergency department visits). Shared decision making between providers and patients to initiate or renew opioid medication should include known general and MS-specific risks and consideration of alternative pain management strategies.

## Data Availability Statement

This research involved analyses of de-identified health care data and cannot be made publicly available.

